# Intersecting Syndemics of Household Water Insecurity and HIV/AIDS: Implications for Dermatological Health among People Living with HIV in Kenya

**DOI:** 10.1101/2025.09.16.25335954

**Authors:** Godfred O. Boateng, Olushina Ayo Junior Ale, Mavis Odei Boateng, Patrick M. Owuor, Ellis Adjei Adams

**Author notes:** Corresponding Author: Godfred O. Boateng, PhD, This work started at the University of Texas at Arlington, Arlington, Texas by Godfred Boateng and completed at York University, North York, Canada.

## Abstract

**Objectives:** In tropical and resource-poor settings where household water insecurity is prevalent, the risk of dermatological conditions is significantly high among People living with HIV (PLHIV). However, no study has examined the nature of this relationship in Kenya. Thus, this study assessed the effect of water insecurity on dermatological conditions among PLHIV in Kenya.

**Methods:** Data for this study were drawn from the Resource Insecurity and Well-being Study with a focus on PLHIV in Kenya (N = 1,132). Data collected included measures of household water insecurity experiences, dermatological conditions, use of improved/unimproved water sources, environmental risk factors, and sociodemographic factors. Following descriptive and bivariate analysis, complementary log-log regression models assessed the effect of water insecurity on each skin condition in three multivariate models. This was followed by predicted probabilities that examined the intersecting syndemics of water insecurity over water source and gender.

**Results:** Of the 1,132 PLHIV, 16.2% reported having skin infections, 16.3% experienced skin itching, and 9.4% experienced skin sores in the last month. In the multivariate models, water insecurity was a significant predictor of skin infections (OR: 1.03; 95% CI: 1.01, 1.04), skin itching (OR: 1.02, 95% CI: 1.01, 1.03) and skin sores (OR: 1.04, 95% CI: 1.02, 1.06). The interaction between water insecurity and improved water sources showed decreased skin sores and itching odds.

**Conclusions:** Household water insecurity is a significant predictor of dermatological conditions among PLHIV. Policies aimed at improving access to clean water and sanitation are essential in promoting their well-being.

## 1. Introduction

Dermatological conditions are estimated to affect 1.8 billion people at any time. These conditions are among the most visible and psychologically burdensome manifestations of HIV/AIDS (1). Approximately 90% of people living with HIV (PLHIV) experience at least one skin condition during their illness, including fungal infections, bacterial dermatoses, viral eruptions, and neoplastic diseases like Kaposi’s sarcoma (2,3). These skin conditions often serve as early clinical indicators of HIV infection and can reflect the degree of immunosuppression (4,5). In resource-limited settings, however, the management of dermatological conditions is frequently deprioritized, even though they significantly affect patient comfort, social stigma, and quality of life (6,7).

The high burden of dermatological diseases in PLHIV is particularly concerning in tropical and low-income regions, where basic hygiene is difficult to maintain due to household water insecurity (8). Water insecurity is a condition when at least one of these variables (affordability, reliability, adequacy, and safety) is significantly reduced or unattainable to threaten or jeopardize well-being, which includes, but is not limited, to physical and mental health and the capacity to undertake necessary productive, social, and cultural activities (9). Water insecurity impairs the ability to bathe, wash clothes or bedding, clean wounds, and maintain sanitary living environments (10). This can contribute to the onset and worsening of various skin diseases, especially those caused by pathogens that thrive in warm, humid, and unsanitary conditions (11). For immunocompromised populations, such as PLHIV, these hygiene constraints can escalate into recurrent or severe skin infections that interfere with overall health and treatment adherence (12).

Kenya has an estimated 1.36 million people living with HIV, with a prevalence rate of 3.3% in 2023, with notable regional disparities (13,14). Simultaneously, only about 36% of the population had access to basic drinking water services, and far fewer, just 34%, had consistent access to clean or safely managed water sources (15). The overlapping challenges of chronic water shortages, inadequate sanitation infrastructure, and the high burden of HIV create a context in which syndemics, interacting epidemics with shared structural drivers, are likely to occur (16). Syndemic theory, first conceptualized by Merrill Singer, highlights how co-occurring epidemics driven by social, environmental, and economic inequalities interact to produce worse health outcomes than each condition would alone (17,18). In the case of HIV/AIDS and household water insecurity, these syndemics do not merely coexist; they amplify each other through biological vulnerability and social disadvantage. For people living with HIV in water-insecure environments, the immune suppression caused by HIV infection interacts with water-related hygiene barriers to increase exposure to pathogens and reduce the body’s ability to resist or recover from infections (17). This interaction can give rise to a cascade of dermatological complications that compromise physical health and social well-being due to the visible nature of many skin diseases.

Despite the considerable burden of dermatological conditions among PLHIV, the intersection between these skin-related health outcomes and structural determinants like water insecurity remains poorly explored. In Kenya, the dual challenges of widespread HIV and chronic household water scarcity create a potent context for what scholars have termed a "syndemic", a set of linked health problems that interact and reinforce each other due to common social and environmental drivers (19). Emerging research supports the relevance of this framework. Studies in western Kenya, where HIV prevalence is high, reveal that water insecurity is strongly associated with adverse HIV-related outcomes. For instance, households experiencing higher levels of water insecurity have significantly greater odds of AIDS-defining illnesses and lack of viral suppression (20). These clinical effects may stem from both biological and behavioural mechanisms, including increased exposure to pathogens, reduced ability to maintain hygiene, and impaired adherence to antiretroviral therapy.

The problem is further compounded by the gendered dynamics of water access. In rural and peri-urban Kenyan communities, women are the primary water collectors. When water becomes scarce or difficult to access, women must travel long distances or make difficult trade-offs, such as reusing unclean water or forgoing hygiene to conserve limited resources (21). These trade-offs can significantly worsen skin conditions in both women and other household members, particularly when shared water is used for multiple domestic and sanitation purposes.

Complicating the situation further is the under-recognition of dermatological conditions as critical components of HIV care. While skin manifestations often precede more serious opportunistic infections and can act as early diagnostic clues, their treatment is frequently marginalized in resource-constrained settings (22). This neglect may arise from competing healthcare priorities, limited training among healthcare providers, or a lack of dermatological medications in public health clinics.

The combination of HIV, water insecurity, and neglected skin health creates a feedback loop. PLHIV with untreated skin infections may avoid social interactions or clinic visits due to stigma, further delaying care. PLHIV who face daily challenges in securing clean water will be severely constrained in following hygiene recommendations or wound-care protocols. Studies show that individuals in water-insecure households report more frequent illness and greater difficulty in maintaining health-seeking routines (23).

Importantly, these burdens are not evenly distributed. Regional disparities in HIV prevalence and water access across Kenya mean that some counties experience more intense health risks than others. For example, counties like Homa Bay and Siaya have some of the highest HIV rates in the country and also face seasonal water shortages that exacerbate household vulnerability (20). These localized intersections offer critical opportunities for targeted interventions, yet they remain underexplored in public health programming. Given this context, the application of syndemic theory becomes useful and essential. It allows policymakers and researchers to move beyond siloed views of water, HIV, or skin health as separate issues. Instead, it frames these problems as mutually reinforcing and tied to broader structural inequalities such as poverty, gender norms, and weak health infrastructure. This perspective encourages holistic solutions, including the integration of water-access programs with HIV care services, and the inclusion of dermatological evaluations in community-based health outreach. Consequently, this paper seeks to advance our understanding of how water insecurity and HIV/AIDS converge to worsen dermatological health among PLHIV in Kenya.

## 2. Methods

### 2.1. Ethics Statement

We received IRB approval from the Institutional Review Boards at the University of Texas at Arlington and the University of Notre Dame (21-05-6613). Also, ethical approval was obtained from the Africa Medical Research Institute (AMREF) Ethics and Scientific Research Committee (ESRC-P1396-2023). Data collection on measures used in this study started July 10, 2023 and ended July 28, 2023. All participants provided verbal informed consent that was documented as part of the data collection process. Minors were not included in this study. We confirm that the study complied with ethical standards outlined in the Belmont Report and/or Declaration of Helsinki

### 2.2. Study setting and population

This study was made possible through a collaboration between the University of Texas at Arlington, York University, the University of Notre Dame and Pamoja Community-Based Organization (CBO) in Kenya. Pamoja CBO, established in 2007, is a grassroots organization that aims to empower communities to identify and address the most significant needs affecting their health and well-being. Among its five thematic areas, the organization runs the Orphans and Vulnerable Children Project, which aims to build the resilience of families and children affected by HIV/AIDS to meet their health, economic, education, and social development needs (24). Consequently, collaborating with Pamoja CBO, we implemented the Resource Insecurity and Well-being Study among PLHIV in the Kisumu West and Seme sub-counties in Kisumu County, where the organization had implemented community HIV prevention interventions for the last 10 years. The area is predominantly rural, with pockets of peri-urban populations near Kisumu City (24).

### 2.3. Study design and data collection

This study aimed to examine how resource insecurity (e.g., household water insecurity) influences health conditions (e.g., dermatological conditions) among PLHIV in Kenya. Data were collected from Seme sub-counties (n=766) and Kisumu West sub-counties (n=366), between August and September of 2023, culminating in a total sample size of 1,132 PLHIV. The surveys were administered using a stratified two-stage cluster sampling, in which each sub-county was considered as a cluster and households were randomly selected based on a manual mapping of where PLHIV were served by the Community Based Organization. Participants were identified from selected households in each of the two sub-counties. Participants were screened for individuals who were most knowledgeable about the different forms of resource insecurities at the household level. Surveys and interviews were limited to participants who were 16 years and older. Data were collected on participants’ socio-demographic characteristics, environmental risk factors, resource insecurity (i.e., food insecurity, water insecurity, energy insecurity, housing insecurity), sources of household resources (e.g., fuel, water, food), dermatological conditions (itching skin, skin sores/ulcers, diverse skin infections), experiences during COVID, and mental health conditions.

### 2.4. Outcome Measures

With dermatological conditions being more prevalent among PLHIV with lower socio-economic standing, three questions informed our outcome measures. Participants were asked disparately if they had experienced skin itching, skin sores/ulcers, and skin infections in the past month. Having a binary response in each case, we coded the responses of the participants to be No (0) and Yes (1).

### 2.5. Key Predictor Variable

The key independent variable was water insecurity, measured using the household water insecurity scale (25). This is a 12-item scale that consists of questions exploring multiple components of water insecurity (accessibility, availability and use). The intensity of water insecurity was assessed with follow-up questions asking whether this condition was experienced never, rarely, sometimes, or often (coded 0,1,2 or 3) with a range of 0-36. The Cronbach’s alpha of the 12 items was 0.97 with an average interitem covariance of 0.59, indicative of the strength of the scale. Higher scores suggest higher experiences of water insecurity, while lower scores are indicative of fewer experiences of water insecurity in the past month.

### 2.6. Covariates

We accounted for biosocial and socio-cultural factors. For biosocial factors, participants were asked for information on age, which was used as a count variable and their gender was coded as male (0) and female (1). Socio-cultural factors included questions on the number of children less than 5 years old in the household, the marital status of the participant coded as single, unmarried (1), separated/divorced/widowed (2), and married/cohabiting (3); neighborhood was coded as Seme sub-county (1) and Kisumu sub-county (2); education was coded as No education (0), Primary (1), Middle/Junior High (2), Secondary/Senior High (3), and Tertiary (4). Participants were also asked to rank their socio-economic status based on a scale of 0-10, with a score of 10 indicative of high socio-economic standing in society. Participants provided information on average monthly income and their perception of their health, ranking their health from very good (1) to very bad (5). Additionally, we coded the primary source of water into improved and unimproved sources following the Joint Monitoring Programme for Water Supply and Sanitation.

### 2.7 Data Analysis

Data analyses were conducted using univariate, bivariate and multivariate regression. We used univariate analysis to assess means and proportions of sample characteristics. We then examined the bivariate relationship between potential confounders, key variables of interest, and the outcome variable. Potential significant confounders at *p*<0.2 were included as covariates in the multivariate model. Due to the asymmetric nature of our binary outcome variables, we resorted to using a generalized linear model with a complementary log-log (cloglog) link function, which is appropriate for modelling rare events and accommodates skewed distributions. In a linear regression model, the expected value of the outcome (dependent) variables, *Y*, is modeled against the set of linear predictors, *X*_1_; *X*_2_;… *X*_k-1_, where the *X*_i_ (i= 1, 2,…, *k*-1) represent the predictor (independent) variable and the covariates to be adjusted for. When the *Y*i (i=1, 2,…, *n*) are independent, identically distributed Bernoulli random variables, the expected value is the proportion of positive (*Y* =1) responses, π, which is also referred to as ‘‘predicted probability.” However, logit and probit models assume that π (x) approaches 0 at the same rate as it approaches 1. The complementary log-log modes assume π (x)= 1 – exp (– exp (α + β_X_)) or equivalently log (− log (1 − π(x))) = α + βx. Following this assumption, the complementary log-log link function was found to be appropriate for modelling the three outcome variables – itchy skin (83.7% vs. 16.3%), skin sores (90.6% vs. 9.7%), and skin infections (83.8% vs 16.2%) that assumed asymmetric distribution. Model performance for each outcome variable was assessed using post-estimation statistics provided by the Fitstat command in Stata 17 (26), including the log likelihood values, Akaike Information Criterion, Bayesian Information Criterion, McFadden’s pseudo-R-squared, and the percentage of correctly classified observations, which serve as appropriate fit indices for generalized linear models. A lower AIC and BIC value and a higher McFadden’s R^2^ associated with the cloglog models make it superior to a logistic model.

Additionally, we generated a margins plot in Stata to graph the predicted probabilities of dermatological conditions for PLHIV at various levels of household water insecurity over gender and water source. This highlights the syndemics of HIV, water insecurity, and their interaction with gender or water source, estimating the compounding probability effect on dermatological conditions.

**Table 1.** Descriptive Statistics of Predictor and Outcome Measures.

## 3. Results

### 3.1. Descriptives

The sample consisted of 1,132 people living with HIV (PLHIV) in Kenya. The mean household water insecurity score was 14.2 (SD = 9.4). The mean age of participants was 44 years (SD = 12.5). Most participants were female (88.1%), while 11.9% were male. On average, participants reported having fewer than one child under five years of age (M = 0.7, SD = 1.5). In terms of marital status, nearly half were either separated, divorced, or widowed (48.9%), followed by those who were married or cohabiting (45.8%) and those who were single (5.4%). Most participants (67.7%) lived in Seme Sub-Counties, and 32.3% were from Kisumu West Sub-Counties. Educational attainment was generally low: 73.6% had completed only primary education, 19.2% had reached middle or junior high school, and smaller proportions had secondary/senior high (1.5%) or tertiary education (1.3%). About 4.4% had no formal education. Subjective socio-economic status averaged 3.0 (SD = 1.3), and average reported monthly income was KSh 5,870, ranging from 0 to 60,000. Perceived health status, indexed as poor health, had a mean score of 1.63 (SD = 0.67). Most respondents relied on unimproved sources of water (82.7%), with only 17.3% accessing improved sources.

### 3.2. Dermatological Conditions

The prevalence of dermatological issues was notable among participants. Skin infections were reported by 16.2% of respondents, 16.3% reported skin itching, and 9.4% reported experiencing skin sores.

### 3.3. Bivariate and Multivariate (Skin Infections)

In the bivariate model, each one-point increase in the household water insecurity scale was associated with a 1.03 times greater likelihood of reporting skin infections (95% CI: 1.01–1.04, p < 0.01). This association remained consistent in the multivariate model, where a one-unit increase in water insecurity scores was associated with being 1.03 times more likely to report skin infections (95% CI: 1.01–1.05, p < 0.001), controlling for all other variables. Female participants had 1.78 times higher odds of reporting skin infections than male participants in the adjusted model (95% CI: 1.04–3.09, p < 0.05). No significant difference by gender was observed in the bivariate model. Participants who were married or cohabiting were 2.45 times more likely to report skin infections than single individuals in the bivariate model (95% CI: 1.00–6.02, p < 0.05), and 3.22 times more likely in the multivariate model (95% CI: 1.22–8.49, p < 0.05). No statistically significant difference was observed for separated, divorced, or widowed individuals. With regards to level of education, respondents with middle or junior high education were 2.88 times more likely to report skin infections than those with no education (95% CI: 1.10–7.58, p < 0.05), and those with tertiary education were 6.65 times more likely (95% CI: 1.65–26.71, p < 0.01), after adjusting for other covariates. These relationships were not statistically significant in the bivariate model. A unit increase in subjective socio-economic status was associated with 0.76 times lower odds of reporting skin infections in the adjusted model (95% CI: 0.67–0.86, p < 0.001), and 0.80 lower odds in the bivariate model (95% CI: 0.70–0.90, p < 0.001). Participants using an improved source of water were 0.60 times less likely to report skin infections compared to those using unimproved sources (95% CI: 0.38–0.94, p < 0.05), in the adjusted model. This association was not significant in the bivariate model. Other factors like age, number of children under five, neighbourhood, health status, and income, were not significantly associated with skin infections in either model. Although monthly income was statistically significant (p < 0.001), an odds ratio of 1.00 indicates no meaningful effect.

The complementary log-log model predicting the presence of skin infections showed improved fit over the intercept-only model, with a log-likelihood of –462.49 versus –498.28. The likelihood ratio test was statistically significant (χ² (15) = 71.60, p < 0.001), indicating that the full model provided a significantly better fit. Model performance was modest, with a McFadden’s R² of 0.072 and a Cragg & Uhler’s R² of 0.105. Efron’s R² was 0.067, and the maximum likelihood R² was 0.062, suggesting satisfactory overall explanatory power. Given the nature of the outcome and the moderate improvement over the null model, the cloglog model was retained for interpretation of covariate effects.

**Table 2:** Bivariate and multivariate complementary log-log regression models showing the relationship between predictor variables and dermatological conditions among PLHIV in Kenya.

### 3.4. Bivariate and Multivariate (Skin Sores)

In the bivariate model, a unit increase in household water insecurity scores was associated with 1.03 higher odds of reporting skin sores (95% CI: 1.01–1.05, p < 0.001). This association remained statistically significant after adjusting for covariates, with a unit increase in water insecurity associated with 1.04 higher odds of reporting skin sores (95% CI: 1.01–1.06, p < 0.001). Married or cohabiting participants were 5.20 times more likely to report skin sores relative to single individuals in the multivariate model (95% CI: 1.39–19.32, p < 0.05), although this association was not statistically significant in the bivariate model. No statistically significant effects were observed for those who were separated, divorced, or widowed. Subjective socio-economic status was inversely associated with the likelihood of skin sores. A unit increase in perceived socio-economic status was associated with 0.68 times lower odds of reporting skin sores in the bivariate model (95% CI: 0.57–0.81, p < 0.001) and 0.66 times lower odds in the adjusted model (95% CI: 0.55–0.77, p < 0.001). Use of an improved water source was associated with a lower likelihood of skin sores. Participants using improved water sources were 0.43 times as likely to report skin sores in the bivariate model (95% CI: 0.22–0.85, p < 0.05) and 0.39 times as likely in the multivariate model (95% CI: 0.20–0.78, p < 0.01). No significant associations were observed for age, gender, number of children under five, neighborhood, health status, or average monthly income in either model.

The complementary log-log model predicting the experience of skin sores showed improved fit over the intercept-only model, with a log-likelihood of −318.05 versus −351.42. The likelihood ratio test was statistically significant (χ²(15) = 66.75, p < 0.001), indicating that the full model provided a significantly better fit. Model performance was modest, with a McFadden’s R² of 0.095 and a Cragg & Uhler’s R² of 0.124. Efron’s R² was 0.062, and the maximum likelihood R² was 0.058, suggesting satisfactory overall explanatory power. Given the nature of the outcome and the moderate improvement over the null model, the cloglog model was retained for interpretation of covariate effects.

### 3.5. Bivariate and Multivariate (Skin Itching)

In the bivariate model, a unit increase in the household water insecurity scores was associated with a1.07 times higher odds of reporting skin itching (95% CI: 1.00–1.13, p < 0.05). This association remained significant in the multivariate model, though slightly attenuated, with each unit increase corresponding to 1.02 higher odds of experiencing skin itching (95% CI: 1.01–1.04, p < 0.01), controlling for all other variables. Participants who were married or cohabiting were 4.49 times more likely to report skin itching relative to single individuals in the bivariate model (95% CI: 1.42–14.16, p < 0.05), and 4.94 times more likely in the multivariate model (95% CI: 1.52–16.05, p < 0.01). No significant differences were observed for separated, divorced, or widowed participants. Although not statistically significant, respondents with tertiary education were 3.62 times more likely to report skin itching compared to those with no education in the multivariate model (95% CI: 0.91–14.34). No significant associations were found for other education levels. Perceived socio-economic status was significantly associated with a lower likelihood of skin itching in the multivariate model. A unit increase in subjective socio-economic status was associated with a 0.86 lower odds of reporting skin itching (95% CI: 0.76–0.99, p < 0.05). This association was not statistically significant in the bivariate model. Use of an improved water source was associated with a lower likelihood of reporting skin itching. Participants using improved sources had 0.49 lower odds of reporting skin itching in the bivariate model (95% CI: 0.30–0.79, p < 0.01), and 0.44 lower odds in the adjusted model (95% CI: 0.26–0.72, p < 0.01). No significant associations were found for age, gender, number of children under five, neighbourhood, income, health status, or other levels of education in either model.

The complementary log-log model predicting the experience of skin itching showed improved fit over the intercept-only model, with a log-likelihood of −479.78 versus −501.56. The likelihood ratio test was statistically significant (χ²(15) = 43.58, *p* < 0.001), indicating that the full model provided a significantly better fit. Model performance was modest, with a McFadden’s R² of 0.043 and a Cragg & Uhler’s R² of 0.064. Efron’s R² was 0.032, and the maximum likelihood R² was 0.038, suggesting limited overall explanatory power. Given the nature of the outcome and the moderate improvement over the null model, the cloglog model was retained for interpretation of covariate effects.

### 3.6. Moderating Effect of Using Improved Water Sources

Following the significant effect of primary source of water with the potential to moderate the effect of household water insecurity, we run interactions between the two variables to assess main and interaction effects, adjusting for covariates. In the skin infections model, only the main effect of water insecurity scores was significant (OR: 1.03, 95% CI: 1.01, 1.05). The main effect of using improved sources of water was not significant; neither was the interaction effect between the two measures, although the results reflected a buffering effect (OR: 0.98, 95%CI: 0.94, 1.02). In the model on skin sores, the main effect of water insecurity (OR: 1.05, 95%CI: 1.02, 1.07) and the interaction effect of water insecurity and improved water source (OR: 0.92, 95%CI: 0.87, 0.98) were statistically significant. Indicating, the use of improved water sources decreased the likelihood of experiencing skin sores. However, the main effect of the improved water was not significant. Similarly, for skin itching, the main effect of water insecurity (OR: 1.03, 95%CI:1.01, 1.04) and interaction of water insecurity and the use of improved water source (0.95, 95%CI: 0.91,0.99) were statistically significant. This signifies that using improved water sources moderated the effect of water insecurity on experiencing skin itching. However, the main effect of using an improved water source was not significant.

### 3.7. Predicted Probabilities

We further assessed interactions using margins for predicted probabilities. We calculated the predicted probability for the interaction of (a) water insecurity and water source, and (b) water insecurity and gender for each of the models. This enabled the examination of the syndemics of HIV, water insecurity, and/or water source or gender.

#### Skin Infections

A predicted probability of 0.12 in Figure 1 suggests there is a 12% chance that PLHIV who score 0 on the water insecurity scale but do not use an improved water source will experience skin infections. This probability increases to 15%, 19%, and 24% when water insecurity scores increase to 15, 21, and 30, respectively. Comparatively, the probability is much lower for those using improved water sources despite their water insecurity scores. At a water insecurity score of 0, 15, 21, and 30, the probability of experiencing skin infections was 8%, 10%, 13%, and 16%, respectively.

**Figure 1.**
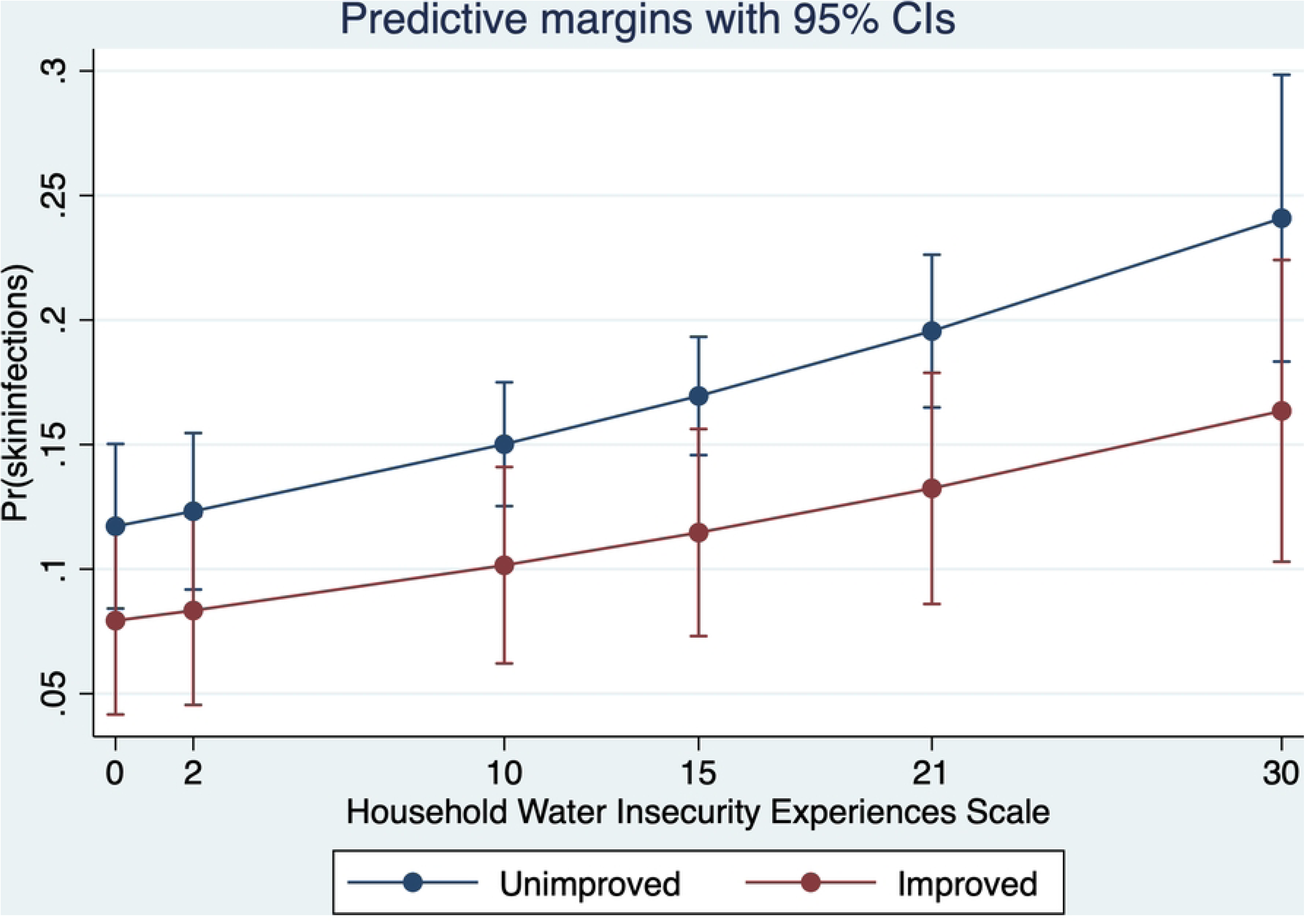
Predicted Probabilities of Skin Infections for Water insecurity over Water source.

With a focus on the interaction between gender and water insecurity (Figure 2), the predicted probability of males experiencing skin infections at water insecurity scores of 0, 15, 21, and 30 correspond to 8%, 12%, 13%, to 17%, respectively. The predicted probabilities for females are significantly higher at the same levels. The probability of experiencing skin infections corresponds to 11% (WI:0), 17% (WI:15), 19% (WI:21), and 24% (WI:30).

**Figure 2.**
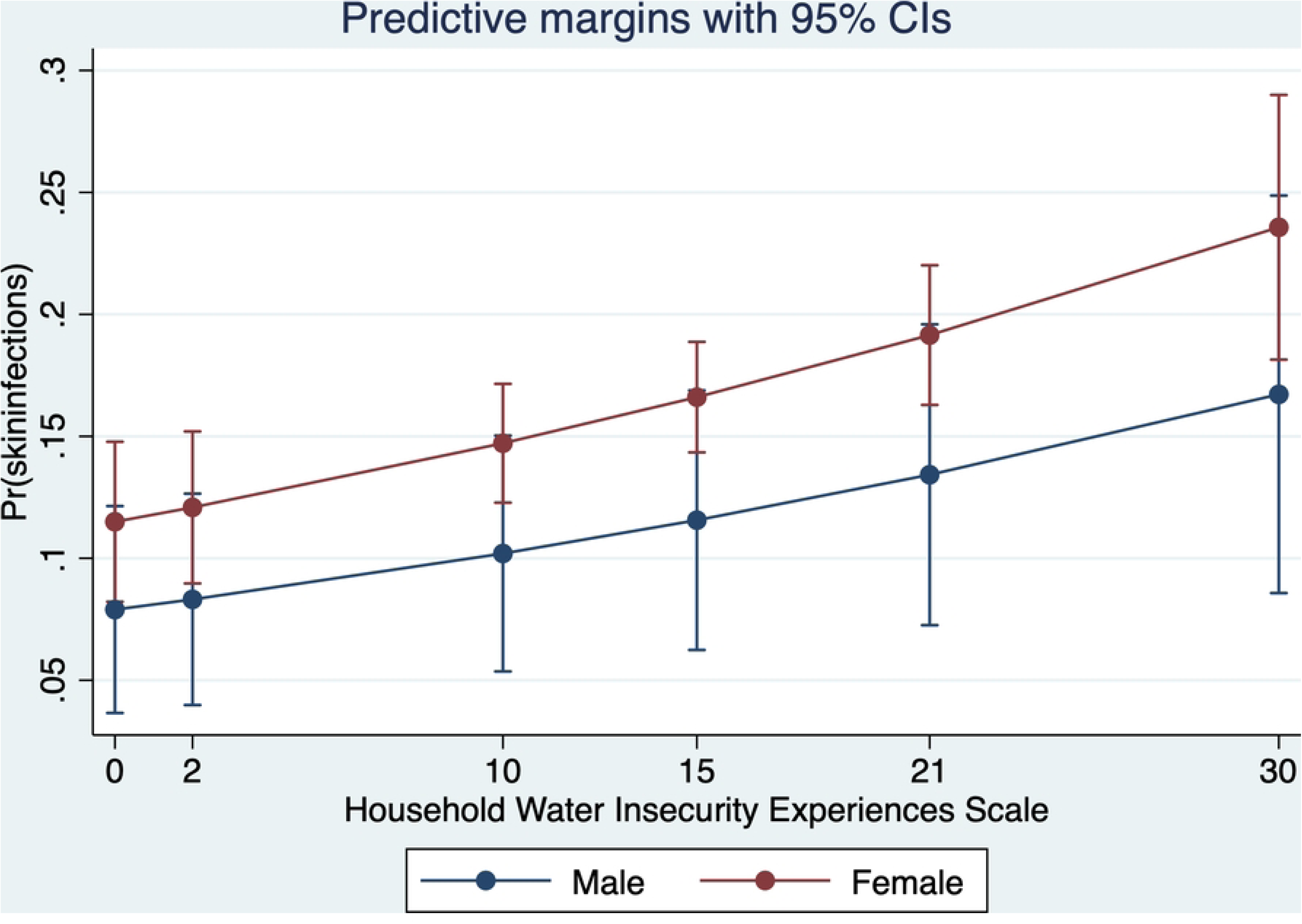
Predicted Probabilities of Skin Infections for Water Insecurity by gender.

#### Skin Sores

The predicted probabilities of experiencing skin sores for the use of unimproved water sources at water insecurity scores of 0, 15, 21,30, start at 6%, 10%, 13%, and 17%, respectively (Figure 3). However, the probability is lower for those using improved water sources at the same water insecurity scores. The probabilities start at 3%, 4%, 5% and 7%, respectively. An examination of the interaction of gender and water insecurity produces a much different story. The probabilities are not significantly different for males and females (Figure 4). At all levels of water insecurity, the probability for males and females starts at 5% (WI:0), 9% (WI:15), 11% (WI:21), and 15% (WI:30).

**Figure 3.**
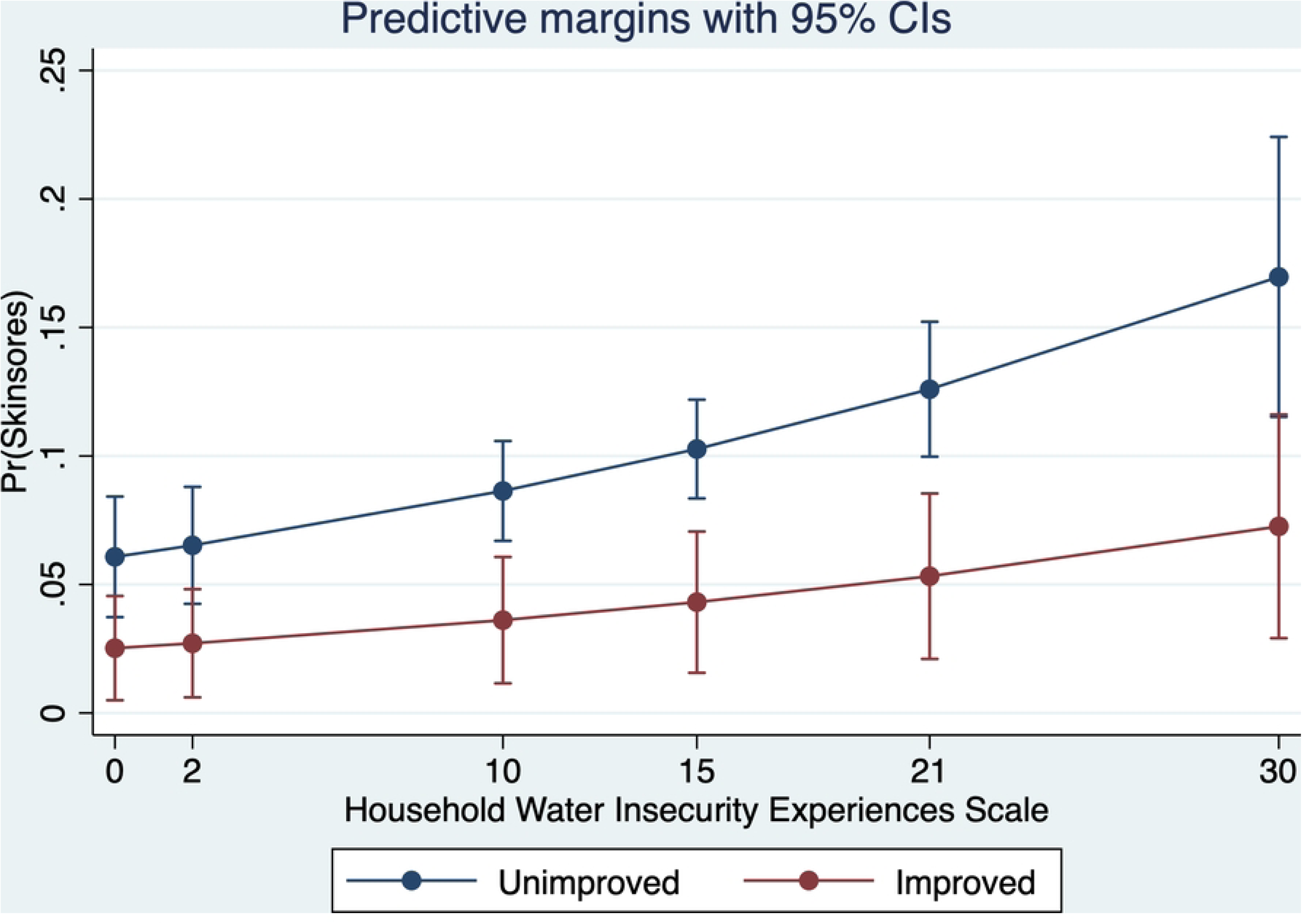
Predicted Probabilities of Skin Sores for Water insecurity over Water source.

**Figure 4.**
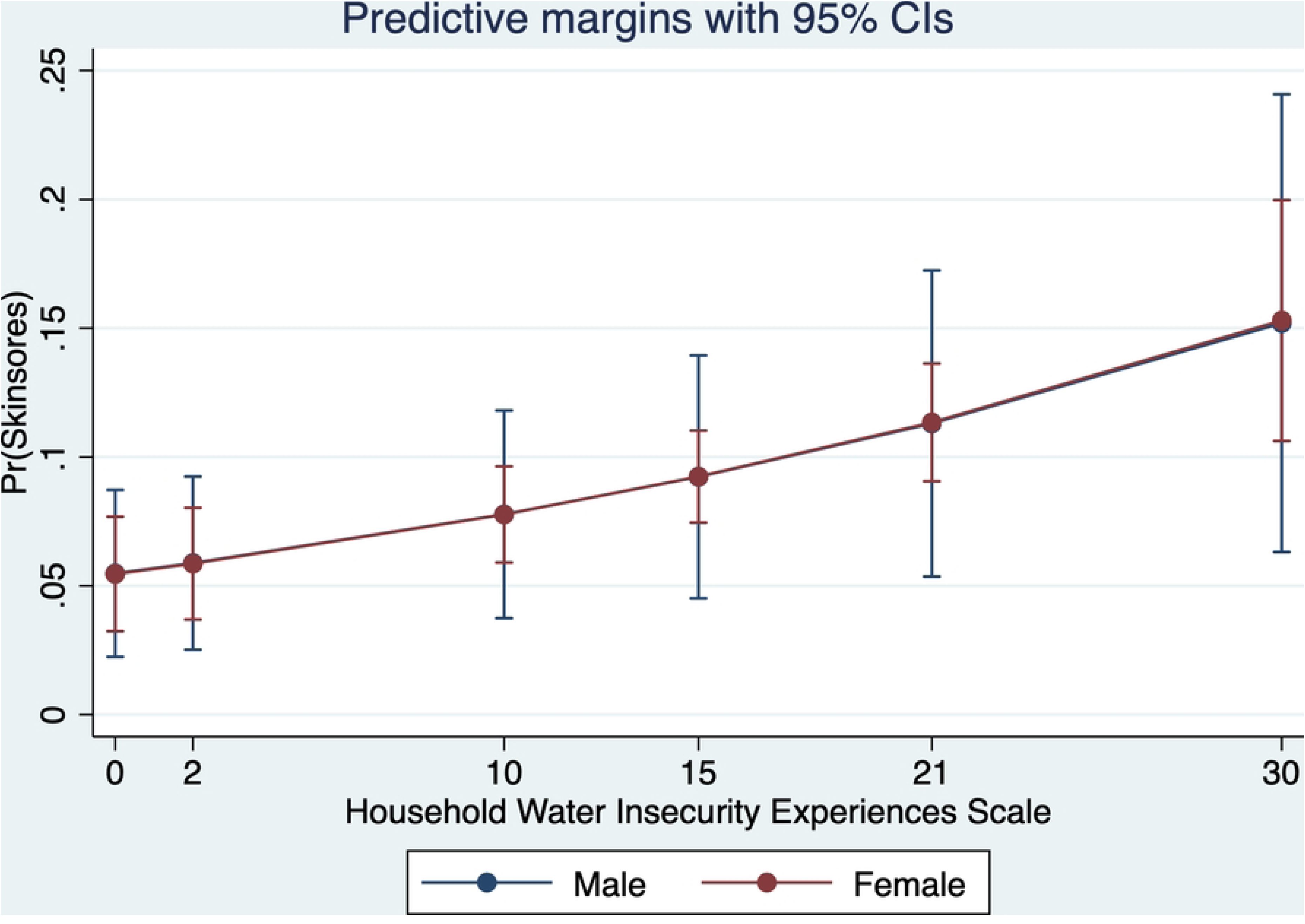
Predicted Probabilities of Skin Sores for Water Insecurity by gender.

#### Skin Itching

The predicted probabilities of experiencing skin itching at water insecurity scores of 0, 15, 21, and 30 correspond to 13%, 18%, 20%, and 24% for those using unimproved water sources. The predicted probabilities were significantly lower for those using improved water sources, 6%, 9%, 10%, and 12%, respectively (Figure 5). An examination of the interaction of gender and water insecurity shows predicted probabilities are slightly higher for females than for males. Examining the same water insecurity scores, the predicted probabilities for males experiencing skin itching were 11%, 15%, 17%, and 20%, respectively. The predicted probabilities for females correspond to 12%, 17%, 19%, and 22% (Figure 6). Thus, among PLHIV, females have a higher predicted probability of experiencing skin itching than males.

**Figure 5.**
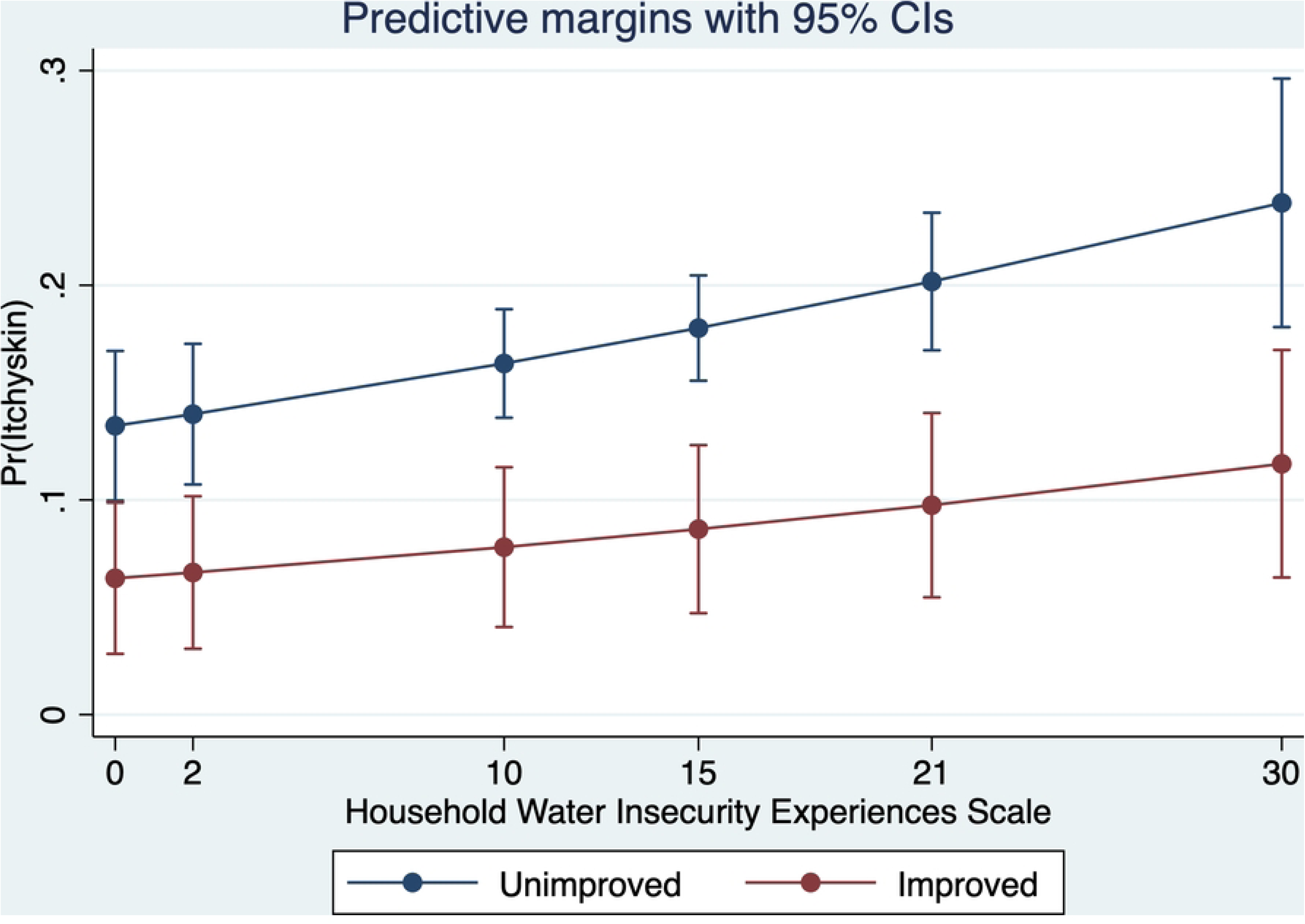
Predicted Probabilities of Skin Itching for Water insecurity over Water source.

**Figure 6.**
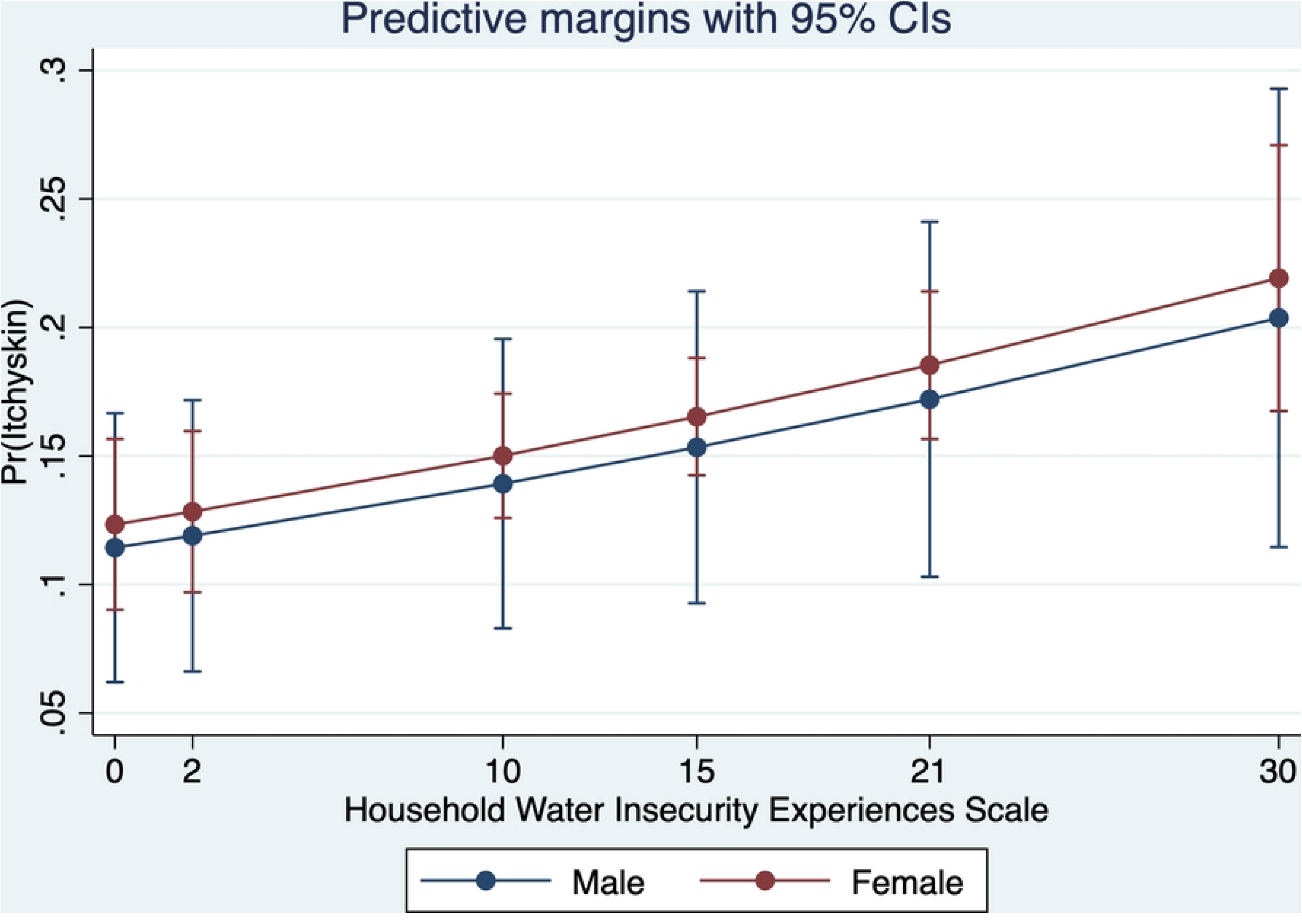
Predicted Probabilities of Skin Itching for Water Insecurity by gender.

## 3. Discussions

This study investigated the relationship between household water insecurity and dermatological conditions, specifically skin infections, skin sores, and skin itching among PLHIV in Kenya. The prevalence of dermatological conditions in this study was substantial: 16.2% reported skin infections, 16.3% experienced skin itching, and 9.4% reported skin sores. These findings show a considerable burden of skin-related morbidity within this population. This is consistent with previous research showing that PLHIV are more susceptible to dermatological conditions due to immune suppression, co-infections, and environmental stressors (27,28).

Most significantly, our study identifies household water insecurity as a key environmental determinant of these conditions. Higher water insecurity scores were consistently associated with increased odds of all three dermatological conditions. These results align with a growing body of literature recognizing water insecurity as a potent and multifaceted stressor affecting both physical and mental health (29–31). Limited access to water may impair basic hygiene practices, increase exposure to contaminated sources, and elevate stress levels, all of which can contribute to compromised skin integrity and infection risk (32,33).

Another significant finding from the study is the moderating effect of the use of improved water sources on the relationship between household water insecurity and both skin sores and itching. Participants using improved water sources had lower predicted probabilities of experiencing these conditions, even when their water insecurity scores were high. This buffering effect reinforces the importance of not only increasing access to water quantity but also improving water quality and safety, a finding echoed in multisite studies on household water insecurity in low- and middle-income countries (33).

Surprisingly, sociodemographic factors played important roles as well. Female participants had significantly higher odds of reporting skin infections than males. This may reflect gendered burdens of caregiving and household water management, which expose women more directly to water-related stressors and poor sanitation (34,35). Marital status was also a predictor, with married or cohabiting individuals more likely to report all three dermatological outcomes possibly due to shared household stress and increased caregiving responsibilities (36).

Interestingly, socio-economic status (SES) emerged as a consistent protective factor. Individuals with higher perceived SES were less likely to report any of the skin conditions, supporting the idea that economic resources help buffer the health impacts of environmental hardship. This aligns with syndemic theory, which emphasizes how intersecting vulnerabilities like poverty and HIV can amplify health risks (37,38).

Furthermore, our findings from the analyses on interactions revealed syndemic patterns. For instance, the probability of skin infections increased sharply with water insecurity, especially among women and those using unimproved water sources. This illustrates the compounded health burden faced by PLHIV experiencing both social and infrastructural vulnerability. While prior research has documented the syndemic interactions between HIV, food insecurity, and mental health (38), our study extends these findings into the domain of physical health, specifically dermatological outcomes, underlining the broader applicability of the syndemic framework.

## 4. Strengths and Limitations

The study’s strengths include its large, community-based sample of PLHIV and its nuanced modeling of interaction effects, which enabled deeper understanding of how water insecurity operates alongside gender and infrastructure factors. However, its cross-sectional design limits causal inference. Self-reported measures of skin conditions may also be subject to reporting bias. Furthermore, while water insecurity was measured at the household level, health outcomes were reported at the individual level, which may introduce measurement mismatch.

## 5. Conclusions

This study adds new evidence to the growing recognition of water insecurity as a critical health determinant for people living with HIV in low-resource settings. We found that household water insecurity is significantly associated with increased risk of skin infections, sores, and itching.

These associations persist even after controlling for demographic and socio-economic factors, underscoring the independent contribution of water-related hardship on dermatological health outcomes. Most significantly, access to improved water source moderated the impact of water insecurity on skin sores and itching, pointing to the protective value of clean water infrastructure. Moreover, gender and socio-economic status emerged as key social factors that shape vulnerability, aligning with syndemic theory, which emphasizes the interaction of structural and biological stressors in producing compounded health burdens.

These findings underscore the need for integrated health and development strategies that address not only HIV care but also the structural determinants of health, especially water access and quality. Efforts to improve water infrastructure, particularly in areas with high HIV prevalence, could play a pivotal role in reducing preventable health complications and improving the quality of life for affected populations.

Future research should explore longitudinal dynamics, investigate other potential health impacts of water insecurity, and test the effectiveness of targeted water interventions in HIV care frameworks. Addressing household water insecurity is not only a matter of development, but also an essential public health policy.

## Data Availability

Data available on request

https://drive.google.com/drive/folders/1uK8E_OXNaOx0RA2tBmkcZFQb8FFDYuie?usp=drive_link

## Declaration of Generative AI in Scientific Writing

During the preparation of this work, the authors did not use AI to generate or write the original draft.

